# Single-cell analyses of CSF and PBMCs from anti-NMDAR encephalitis patients reveals distinct characteristics of T cell subpopulations

**DOI:** 10.1101/2023.07.27.23292878

**Authors:** Sisi Li, Xiang Hu, Yi Yang, Jierui Wang, Zhen Hong, Dong Zhou, Jinmei Li

## Abstract

**Background:** Anti-N-methyl-D-aspartate receptor encephalitis (NMDAR-E) is a common and severe antibody-mediated autoimmune encephalitis. While the roles of B cells and NMDAR antibodies in NMDAR-E have been extensively studied, the involvement of T cell subpopulations in the disease progression remains unclear.

**Methods:** This study conducted single-cell RNA sequencing, single-cell TCR sequencing, and flow cytometry to analyze the T cell subpopulations and their transcriptomic characteristics in NMDAR-E patients and control individuals. Furthermore, it explored the interaction between CD8^+^T cells and B cells through in vitro cell co-culture and cell communication analysis.

**Results:** The study found activated CD8^+^T cell subpopulations in the cerebrospinal fluid (CSF) and peripheral blood mononuclear cells (PBMCs) of NMDAR-E patients, with some subpopulations exhibiting significant TCR clonal expansion. Differential expression gene analysis revealed upregulation of genes related to cytotoxicity, tissue residency, Th1, IFN, or TCR signaling in certain activated CD8^+^T cell and CD4^+^ memory T cell subpopulations. In vitro co-culture experiments demonstrated that CD8^+^T cells from the PBMCs of NMDAR-E patients could induce apoptosis of their own B cells. Cell interaction analysis revealed the existence of interactions between KIR^+^CD8^+^T cells and B cell subpopulations in NMDAR-E patients.

**Conclusion:** This study explored the changes and transcriptomic characteristics of activated CD8^+^T cell subpopulations in the CSF and PBMCs of NMDAR-E patients. Additionally, it discovered the impact of CD8^+^T cells from NMDAR-E patients on their own B cells, providing new evidence for the interaction between CD8^+^T cells and B cells.

## 1. Introduction

Anti-N-methyl-D-aspartate receptor (NMDAR) encephalitis (NMDAR-E) is a recently discovered common autoimmune encephalitis (AE). It is caused by autoantibodies against the NMDAR NR1 subunit, leading to severe diffuse brain inflammation symptoms ^1, 2^, including psychiatric symptoms, cognitive impairments, seizures, complex movement disorders, or unexplained coma ^3, 4^. Current first-line immune therapies include steroids, plasma exchange, and intravenous immunoglobulin, but approximately 20% of patients do not respond to these treatments ^5-7^. Additionally, about 75% of patients require intensive care unit treatment ^3, 8^, and 20-25% are at risk of relapse ^7, 8^. Due to the pathogenicity of NR1-IgG being established ^9-11^, the clinical use of anti-CD20 monoclonal antibodies (such as rituximab (RTX) and ocrelizumab (OFA)) in NMDAR antibody encephalitis is increasingly widespread, though their effectiveness is limited in some patients ^12^. Furthermore, nearly all patients experience long-term sequelae, such as persistent cognitive deficits ^13^. Despite extensive research into the pathogenic role of NR1-IgG autoantibodies in NMDAR-E, previous studies have primarily focused on B cell subsets, such as antibody-producing cells, with limited understanding of other immune cell subsets.

Recent studies have shown that T cell subsets may play a role in the pathogenesis of NMDAR-E. Studies have demonstrated significantly elevated levels of T cell-related cytokines in the cerebrospinal fluid (CSF) of NMDAR-E patients ^14-16^. In brain tissue samples from these patients, infiltration of T cells around brain parenchyma and perivascular spaces has been observed ^17-19^. Animal models of NMDAR-E have revealed infiltration of CD4^+^ and CD8^+^T cell subsets in critical brain regions ^20^. In a mouse model of autoimmune encephalitis induced by repeated streptococcal infections, Th17 lymphocytes play a crucial role in the entry of autoantibodies into the central nervous system (CNS), sustained activation of microglia, and neurological deficits ^21^. Another study established a humanized mouse model by transferring peripheral blood mononuclear cells (PBMCs) from NMDAR-E patients, showing infiltration of CD8^+^T cells in the hippocampal region ^22^. However, despite the observed infiltration of CD8^+^T cells in brain tissues of NMDAR-E patients and animal models, no signs of neuronal apoptosis or necrosis have been found ^17, 19^, suggesting that CD8^+^T cells may not attack neurons through cytotoxic effects. Further exploration is needed to investigate whether CD8^+^T cells play other roles in NMDAR-E.

CSF is a transparent fluid that envelops and protects the CNS, forming a unique local immune environment ^23^. Under healthy conditions, the non-cellular components of CSF primarily consist of serum ultrafiltrate ^23^. Clinically, CSF aids in the diagnosis of CNS inflammatory and degenerative diseases. However, the concentration of cells in CSF is approximately 1,000 times lower than in blood, and the volume that can be safely sampled from each patient is limited. Single-cell transcriptomics is a transformative and rapidly advancing technology primarily used to redefine the heterogeneity of complex tissues in healthy rodents or humans ^24, 25^. However, its application in autoimmune encephalitis such as NMDAR-E is not yet widespread. To our knowledge, only three studies have reported the transcriptomic and immunological characteristics of B cell subsets in the CSF, peripheral blood (PB), and teratoma tissues of NMDAR-E patients ^26-28^. To date, no studies have reported the transcriptomic and immunological characteristics of T cell subsets in CSF and PB of NMDAR-E patients.

Based on this, our study utilizes a combination of single-cell RNA sequencing (scRNA-seq), single-cell T cell receptor sequencing (scTCR-seq), flow cytometry, and in vitro co-culture methods to explore the phenotypes of T cells, TCR clonality, and the interactions between T cell subsets and B cells in NMDAR-E patients (Fig. 1A). This research aims to fill the current research gap regarding T cell subsets in NMDAR-E and provide new insights into the interactions between T cells and B cells.

**Figure 1.**
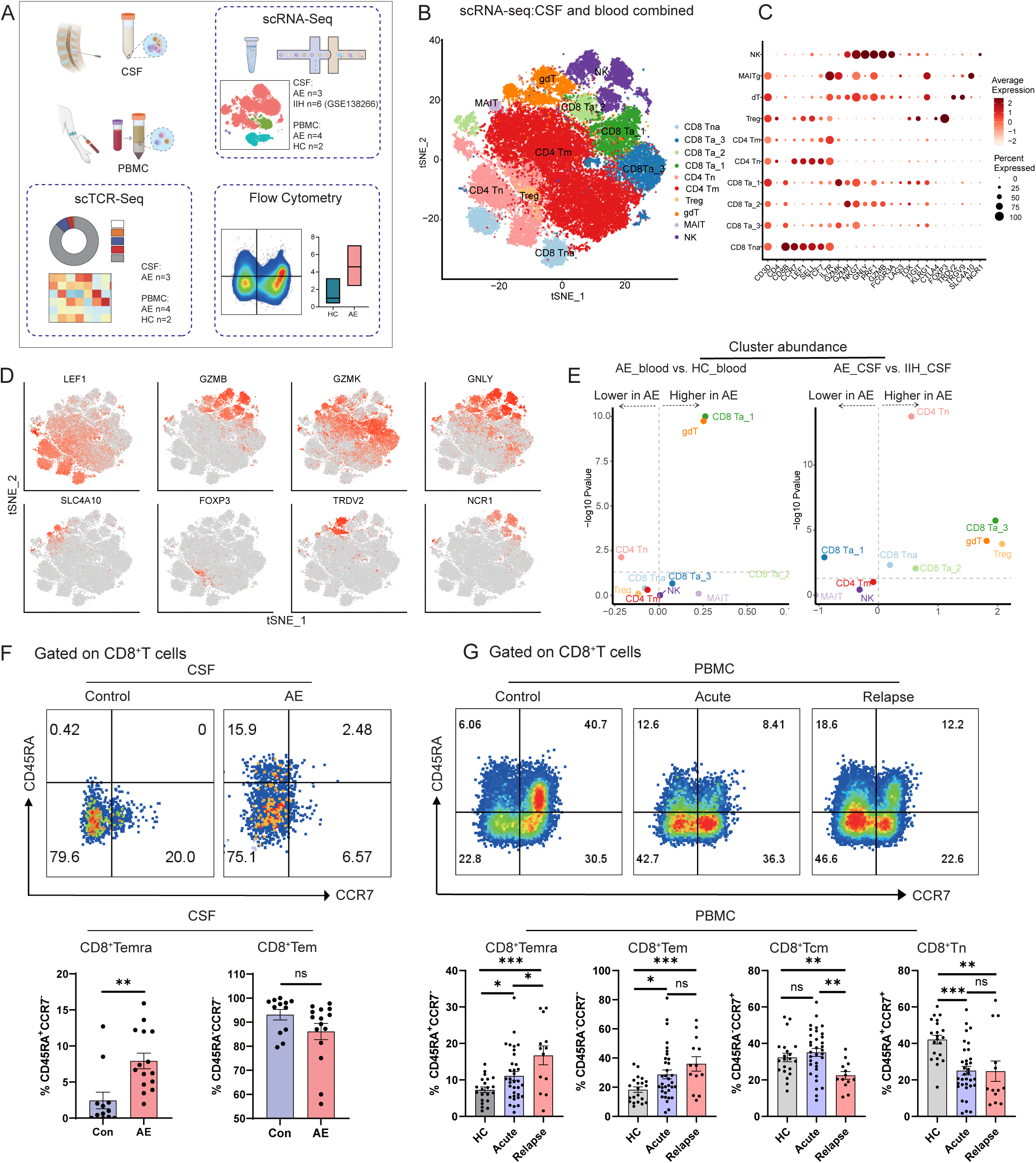
Single-cell transcriptomics analysis the T cell composition of cerebrospinal fluid (CSF) and periphery blood (PB). (A) Schematic representation of the study design. (B) The t-SNE plot displays 10 distinct clusters of cells, each color-coded and identified based on merged single-cell transcriptomes obtained from PB (NMDAR-E, n=4; HC, n=2) and CSF (NMDAR-E, n=3; IIH, n=6) samples. The assigned cluster names are denoted manually. (C) Expression of marker genes in T and NK cells was used to classify subclusters. The dot size indicates the percentage of cells expressing the gene, while color represents the average gene expression level per cell. (D). The t-SNE plot displayed the distribution of selected marker genes within T/NK cell subpopulation. (E) These volcano plots illustrate the comparison of CSF and PB cell subpopulations between NMDAR-E patients and controls. The plots are generated using a beta-binomial regression model and are presented as a fold change (log10) against p value (-log10). The horizontal line denotes the significance threshold. (F) Representative contour plots and summary scatter plots show the percentage of CD45RA^+^CCR7^-^ (CD8^+^Temra) and CD45RA^-^CCR7^-^ (CD8^+^Tem) cells in CSF from control individuals (n = 12) and NMDAR-E patients (n = 15). **p < 0.01, two-tailed unpaired Student’s t test. (G) Representative contour plots and summary scatter plots show the percentage of CD8^+^Temra, CD45RA^+^CCR7^+^(CD8^+^Tn), CD45RA^-^CCR7^+^ (CD8^+^Tcm), and CD8^+^Tem cells in PB from control individuals (n =21), acute phage NMDAR-E patients (n = 34), and relapse phage patients (n=13). **p < 0.01, ***p < 0.001, One-Way ANOVA test. Flow charts of for (F-G) are provided in Figure e-3.

## 2. Methods

### 2.1 Selection of Patients and Controls

All samples were collected from West China Hospital, Sichuan University between January 2021 and January 2023, and recruited patients met the diagnostic criteria for NMDAR-E^1^. The inclusion criteria for NMDAR-E in patients were consistent with our team’s previous report^29^. This study obtained ethical approval from the Institutional Review Board (IRB) of West China Hospital (Protocol No. ChiCTR1900027074) and followed the ethical principles outlined in the Declaration of Helsinki. Exclusion criteria included: (1) patients with coexisting autoantibodies; (2) patients who have received late-stage immunotherapy (such as Rituximab, OFA, cyclophosphamide, etc.); (3) patients who refused to participate.

Inclusion criteria for the CSF control group samples are as follows: (1) Negative results in CSF examination, including routine, biochemical, and IgG tests, as well as negative results for bacterial, fungal and viral tests; (2) Negativity for autoantibodies in the CSF, demyelinating antibodies, and antibodies associated with paraneoplastic syndromes; (3) No Signs of inflammation should be observed in the cranial magnetic resonance imaging (MRI); (4) Patients should be in good physical health; (5) The final diagnosis should be idiopathic intracranial hypertension, chronic headache, or other non-organic central nervous system diseases. The PBMCs control group samples are obtained from healthy adult volunteers matched for age and gender with the enrolled patients.

### 2.2 Sample information for single cell analysis and flow cytometry

This study investigated scRNA-seq and scTCR-seq analyses using samples from nine subjects, consisting of three CSF samples from NMDAR-E patients, four PBMC samples from NMDAR-E patients, and two PBMC samples from control individuals. In addition, six CSF control samples from idiopathic intracranial hypertension (IIH) were obtained from the public dataset GSE138266 and were used for scRNA-seq analysis (Table S1).

Flow cytometry was used to detect changes in T-cell subsets in the CSF and PBMC of patients with NMDAR-E as well as in the controls. The following analyses were conducted: (1) a total of 25 CSF samples (NMDAR-E, n=15; control, n=12) (Table S2) and 55 PBMCs (acute-phase NMDAR-E, n=34; relapse-phase NMDAR-E, n=13; controls, n=21) (Table S3); (2) KIR^+^CD8^+^T cell subsets were analyzed in samples from 40 NMDAR-E (acute severe phase, n=9; acute mild/moderate phase, n=10; relapse-phase, n=13; remission-phase, n=8) as well as 11 controls (Table S4). Due to PBMCs samples separation into two batches for testing, the first batch only examined the CD8^+^ and CD4^+^T cell subsets of the acute-phase and control group samples. In the second batch (as shown in Table S4), the CD8^+^ and CD4^+^T cell subsets of the acute-phase, relapse-phase, and control group samples were simultaneously tested for KIR^+^CD8^+^T cell subset. The analysis in Table S3 includes data from both the first and second batches.

Disease severity was assessed using the mRS scores ^30^, where a score of <3 indicated mild illness and ≥3 indicated moderate/severe illness. Relapse was defined as the recurrence or emergence of new symptoms after a period of at least 2 months marked by clinical stabilization or improvement ^31^.

### 2.3 Collection and processing of CSF and PB samples

PB samples (4-5 mL) were obtained from both NMDAR-E patients and control groups. To isolate PBMCs, Ficoll-Paque medium (Ficoll Paque Plus, GE Healthcare) was used for density gradient centrifugation. After separation, PBMC samples were preserved by freezing them overnight at -80°C and subsequently transferring them to liquid nitrogen for long-term storage. These samples were later used for scRNA-Seq, scTCR-seq, and flow cytometry analysis.

We collected CSF samples from both NMDAR-E patients and control groups via lumbar puncture. Fresh CSF samples of 15-30 mL were required for scRNA-seq and scTCR-seq analysis, while 5-10 mL of fresh CSF samples were needed for flow cytometry analysis. Sample processing was conducted within 2 hours after collection. Fresh CSF samples were used for scRNA-Seq, scTCR-seq, and flow cytometry analysis.

### 2.4 Flow cytometry

PBMCs and CSF cells were suspended in phosphate-buffered saline (PBS) supplemented with 10% fetal calf serum (FCS). The cells were then treated for 30min with a mixture of antibodies. To characterize T cell subsets in CSF and PBMCs, the panel included the following antibodies: CD3-BV605, CD8-APC-Cy7, CD4-PerCP-Cy5-5, CD45RA-BV786, and CCR7-FITC. To analyze KIR^+^CD8^+^T cells in PB, the following antibodies were used: CD3-BV605, CD8-APC-Cy7, CD45RA-BV786, CCR7-FITC, CD158b-PerCP-Cy5-5, and CD158e-PE. For the separation of CD8^+^T cells and B cells, the following antibodies were employed: CD3-BV605, CD8-APC-Cy7, 7AAD-PerCP-Cy5-5, and CD19-BV785. In vitro co-culture of CD8^+^T cells and B cells, as well as measurement of B cell apoptosis, were detected using the following antibodies: CD3-BV605, CD19-BV785, CD38-APC, CD27-PE-Cy7, IgD-APC-Cy7, AnnexinV-PE, and 7AAD-PerCp-Cy5-5. For each experiment, control samples were stained separately and left unstained. Flow cytometry experiments were performed using the LSRFortessa flow cytometer from BD Biosciences. A compensation matrix was developed to facilitate analysis using FlowJo software (v. 10.5.3, FlowJo LLC, BD). Detailed information for each flow cytometry antibody can be found in Table S5.

### 2.5 Single-cell RNA-Seq data analysis

#### Data processing, quality control, and batch correction strategies

The raw data underwent processing utilizing CellRanger software (version 7.0.0) and aligned with the GRCh38 human reference genome. The gene-barcode matrix was analyzed using Seurat package in R (version 4.2.0). During the quality control process, cells with less than 200 detected genes for environmental RNA, cells with mitochondrial gene expression total exceeding 15% for dead cells, and doublets identified by Scrublet (Python 3.7.3) were excluded. In order to mitigate the impact of batch effects, the single-cell variational inference (scVI) method was employed to integrate expression data from different samples ^32^. ScVI is a powerful deep generation model that can learn the transcriptional state of a single cell and implicitly correct the batch effect ^33^.

#### Identification of cell cluster

To perform principal component analysis (PCA), we utilized Seurat’s RunPCA function to select the top 2000 features displaying the most significant expression differences between cells. Subsequently, cell clustering was performed on the top 30 dimensions using the FindClusters function. The resulting clusters were visualized using t-distributed stochastic neighbor embedding (t-SNE), and the identification of cell types within each cluster was determined based on characteristic marker genes.

#### Cell clustering analysis

For the T/NK cell subclustering analysis, we utilized t-tests and beta-binomial generalized linear models from the aod::betabin package. These methods allowed us to examine differences in cluster abundance, measured as cell counts, between NMDAR-E encephalitis and control donors^33^.

#### DEGs and pathway analysis

To identify genes exhibiting differential expression (DEGs), we employed Seurat’s "FindMarkers" or "FindAllMarkers" functions. In addition, we utilized the Wilcoxon test to assess the significance of gene expression differences. Genes expressed in at least 25% of the cells within the target cluster were considered, and a p-value threshold below 0.05 was used to determine statistical significance. To visualize the expression patterns of specific genes, we generated violin plots using the Seurat package. Furthermore, for the enrichment analysis of the identified DEGs in the Kyoto Encyclopedia of Genes and Genomes (KEGG), we utilized the singleseqgset package (https://github.com/arc85/singleseqgset).

#### Cell communication

To investigate the intercellular interactions between KIR^+^CD8^+^T cell and B cell clusters in NMDAR-E patients and controls, we employed the R package CellChat. This package facilitated the identification and visualization of intercellular communication by leveraging known ligand-receptor pairs ^34^. The CellChat package can be accessed at https://github.com/sqjin/CellChat. To explore the interactions between potential ligands and receptors, we analyzed the relative expression levels across different cell types using CellPhoneDB (v3.0.1) (www.CellPhoneDB.org). Heatmaps and bubble plots were generated using the ktplots (v1.2.1) R package to visualize these interactions.

### 2.6 Analysis of single-cell TCR sequencing data

The study utilized Cell Ranger software (version 6.0.1) to identify CDR3 sequences and rearranged TCR genes in the 10x Genomics dataset. The α-β pairing within the framework was considered to determine the dominant TCRs and unique clone types. Clonal populations were denoted by the presence of a clone type in at least two cells, and clonality was determined by the number of cells with the same dominant α-β pairing. Clonotypes were divided into three groups based on the number of T cells expressing the same TCR: ">=3", "2", and "1". Following a similar approach as described in previous studies^35, 36^, we employed a statistical method to assess the sharing of TCR clones based on TCR clonotypes between different clusters. Specifically, to evaluate the sharing of TCR clones in both the NMDAR-E patient CSF and PB datasets, we generated a table for each pair of clusters and computed the number of clone types shared among cells within the same cluster or across different clusters.

### 2.7 In vitro experiments

The PBMCs of NMDAR-E patients and controls were prepared, and CD8^+^T cells and B cells were sorted by flow cytometry and co-cultured at a ratio of CD8^+^T: B cells (5:1). After 1day and 7 days of cultivation, flow cytometry antibody staining was performed, incubated at 4_ in the dark for half an hour, then washed, resuspended, and filtered into flow tubes. The LSRFortessa flow cytometer (BD Biosciences) was used for detection.

### 2.8 Statistical analysis

Descriptive analysis was performed to examine the sociodemographic and clinical data, and the results were expressed as percentages or means using IBM SPSS Statistics version 22.0 (IBM Corp., Armonk, NY, USA). The flow cytometry analysis was conducted using FlowJo software (v. 10.5.3, FlowJo LLC, BD), and the graphs were generated using GraphPad Prism (V.9.0). To assess the differences between two cell subpopulations, an unpaired t-test was employed, while the differences among multiple groups were evaluated using the One-Way ANOVA test. To facilitate image layout, we utilized the commercial software Adobe Illustrator 2020. We described sample sizes and statistical tests methods in the figure legends to provide a comprehensive understanding of the results. Unless otherwise specified, P-values or adjusted P-values less than 0.05 were considered significant.

## 3. Results

### 3.1 Composition and signature marker genes of T/NK cell subsets

After conducting quality control and technical noise adjustment, a total of 45,088 single-cell transcriptomic profiles were included for analysis (Table S6). Additionally, we incorporated 14,536 CSF cells from six idiopathic intracranial hypertension (IIH) patients obtained from a publicly available database as control samples (GSE138266). Detailed gene expression levels for each group can be found in Figure S1A. Through the utilization of t-SNE and assessment of the expression of characteristic marker genes, we thereby classified 59,624 single cells into eight immune cell clusters (Figure S1B-S1D). Our previous research had elucidated B cell phenotypes in the CSF and PB of NMDAR-E patients ^28^. This study primarily focuses on investigating the composition of T cell subpopulations and transcriptomic changes.

In this study, the total number of T/NK cells analyzed from the CSF and PB of NMDAR-E patients and controls was 47,922 (Figure 1B). To classify different T/NK cell subsets, we used marker genes and identified distinct clusters (Figure 1C and 1D). CD4^+^T cells were divided into two groups. Naïve CD4^+^T cells (CD4^+^Tn) exhibited high expression of *CCR7*, *SELL*, and *LEF1* genes, while memory CD4^+^T cells (CD4^+^Tm) demonstrated low expression of these genes. CD8^+^T cell subsets were divided into non-activated CD8^+^T (CD8^+^Tna) cells and activated CD8^+^T (CD8^+^Ta) cells. CD8^+^Tna cells exhibited high expression of *CCR7*, *SELL*, and *LEF1*. CD8^+^Ta cells consisted of three groups, where the first (CD8^+^Ta_1) and third (CD8^+^Ta_3) groups highly expressed *CD8B* and *GZMK* genes and were biased towards the characteristics of effector memory CD8^+^T (CD8^+^Tem) cells, while the second group (CD8^+^Ta_2) highly expressed *GZMH*, *GNLY*, *PRF1*, *GZMB* genes and exhibited similar features to that of effector CD8^+^T cells ^37^. In addition, we also identified gdT (*TRDV2* and *TRGV9*), Treg (*Foxp3* and *CTLA4*), MAIT (*SLC4A10*), and NK (*GNLY* and *NKG7*) cell subsets ^33, 37^.

The proportion of T cell subsets in the CSF and PB differs in NMDAR-E patients. In PB, CD4^+^Tn, CD8^+^Tna, and NK cells are predominant, while in CSF, CD4^+^Tm and CD8^+^Ta cells are predominantly found (Figure S1E). In NMDAR-E patients, the proportion of the CD8^+^Ta_1 subset is higher in PB than in the control group, whereas the proportions of CD8^+^Ta_2 and CD8^+^Ta_3 cell subsets are higher in CSF compared to the control group (Figure 1E). Additionally, we conducted flow cytometry analysis on CD8^+^T and CD4^+^T cell subsets in the CSF and PBMCs of patients with NMDAR-E and control subjects (Figure S2A). The results revealed a significant increase in the number of CD45RA^+^effector memory CD8^+^T cells (CD8^+^Temra) in the CSF of NMDAR-E patients (Figure 1F). In PB, the levels of CD8^+^Temra and CD8^+^Tem cells were both significantly higher in the acute-phase and relapse-phase NMDAR-E patients compared to the control group (Figure 1G). There were no statistically significant differences in the levels of CD4^+^T cell subsets between the two groups in PB and CSF (Figure S2B and S2C).

### 3.2 Transcriptome characteristics of T cell subsets

Through the analysis of the differential expression genes (DEGs) of T cell subpopulations in NMDAR-E patients and controls, the results showed that the CD8^+^Ta cell subpopulation in the CSF and PB of NMDAR-E patients upregulated DEGs related to cytotoxicity compared to the control group (mainly *GZMK*, *PRF1*, and *GZMA* in CSF, and *GZMH* and *NKG7* in PB) ^38, 39^ (Figure 2). The CD8^+^Ta_1, CD8^+^Ta_2, and CD4^+^Tm cell subpopulations in NMDAR-E CSF upregulated DEGs related to tissue residence (such as *CD69*, *CXCR6*, *PRDM1*, *LGALS3*) ^38^ (Figure 2A and 2B). In addition, compared to the control group, CD4^+^Tm, CD8^+^Ta, and gdT cells in the CSF and PB of NMDAR-E patients were found to upregulate DEGs related to co-inhibitory (such as *CTLA4*, *LAG3*, *ANXA1*), Th1-related genes (such as *TBX21*, *CXCR3*, *RUNX3*), interferon (IFN)-related genes (such as *IFNGR1*, *IRF7*, *STAT1*), as well as T cell receptor (TCR) signaling pathway (such as *DUSP2*, *GIMAP1*, *GIMAP5*, *PIK3R1*) ^36^ (Figure 2). Furthermore, this difference was more pronounced in the CSF of NMDAR-E patients.

**Figure 2.**
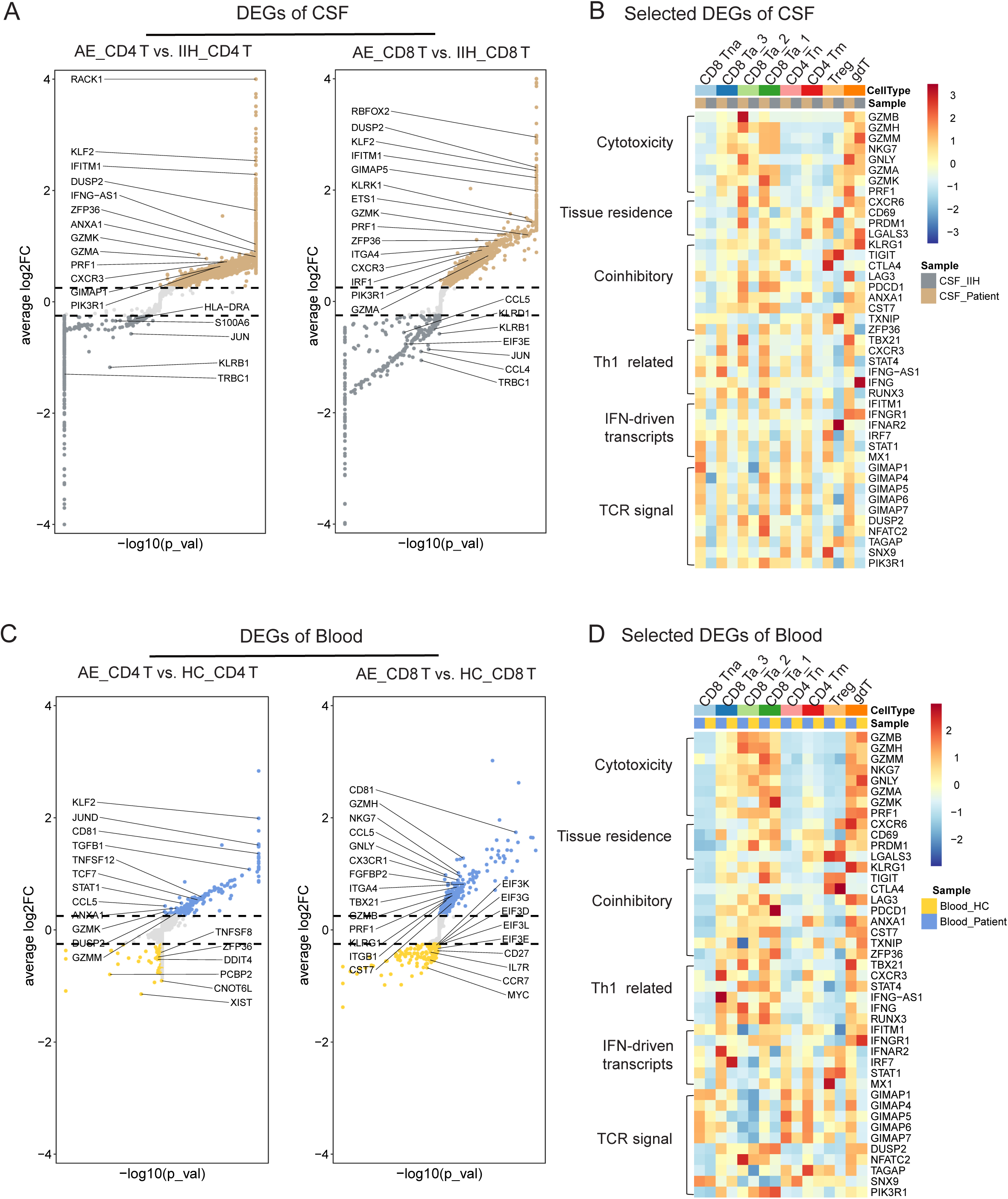
Transcriptomic characteristics of T cell subsets in the CSF and PB of patients with NMDAR-E patients and controls. (A) Differential gene expression (DEG) of CD4^+^T cells and CD8^+^T cells in the CSF of NMDAR-E patients compared to controls. Source data are provided in Table S7. (B) DEGs within T cell subclusters in the CSF of both NMDAR-E patients and control individuals, and calculated the averaged scaled expression levels across all cells within each group. (C) DEG of CD4^+^T cells and CD8^+^T cells in the PB of NMDAR-E patients compared to controls. Source data for are provided in Table S8. (D) DEGs within T cell subclusters in the PB of both NMDAR-E patients and control individuals, and calculated the averaged scaled expression levels across all cells within each group.

### 3.3 TCR clonal characteristics of T cell subsets

Through single-cell TCR sequencing, we observed that compared to the control group, NMDAR-E patients had a higher proportion of T cells with TCR clonotypes in PB, and this phenomenon was more pronounced in the patients’ CSF (Figure 3A and 3B). T cell subpopulations with high TCR clonality (TCR clonotypes ≥3) were mainly distributed in CD8^+^Ta_1, CD8^+^Ta_2, CD4^+^Tm, and MAIT cell subgroups, while the TCR clonality of CD4^+^Tn and CD8^+^Tn cells showed no pronounced changes (Figure 3C-3E). In the CSF of NMDAR-E patients, T cells with TCR clonality showed upregulation of cytotoxicity-related genes (such as *GZMA*, *GZMK*) ^39^ and cell migration-related genes (such as *CCL5*, *CCL4*, *CCL4L2*, *ITGB1*) ^40^, while genes related to naïve/memory status (such as *CCR7*, *SELL*) were downregulated (Figure 3G). Similarly, T cells with TCR clonality in the PB of NMDAR-E patients showed upregulation of DEGs related to cytotoxicity (such as *GZMH*, *GZHB*, *PRF1*) and cell migration (such as *CX3CR1*, *ITGB1*) ^40^ (Figure 3F and 3H). Moreover, we identified numerous shared TCR clonotypes in T cell subpopulations in the CSF and PB of NMDAR-E patients (Figure 3I and 3J). For instance, shared TCR sequences were observed between CD4^+^Tn and CD8^+^Tn in PB, and between CD4^+^Tn and CD4^+^Tm, as well as between CD4^+^Tm and CD8^+^Tem in the CSF, indicating a common clonal origin among these subpopulations.

**Figure 3.**
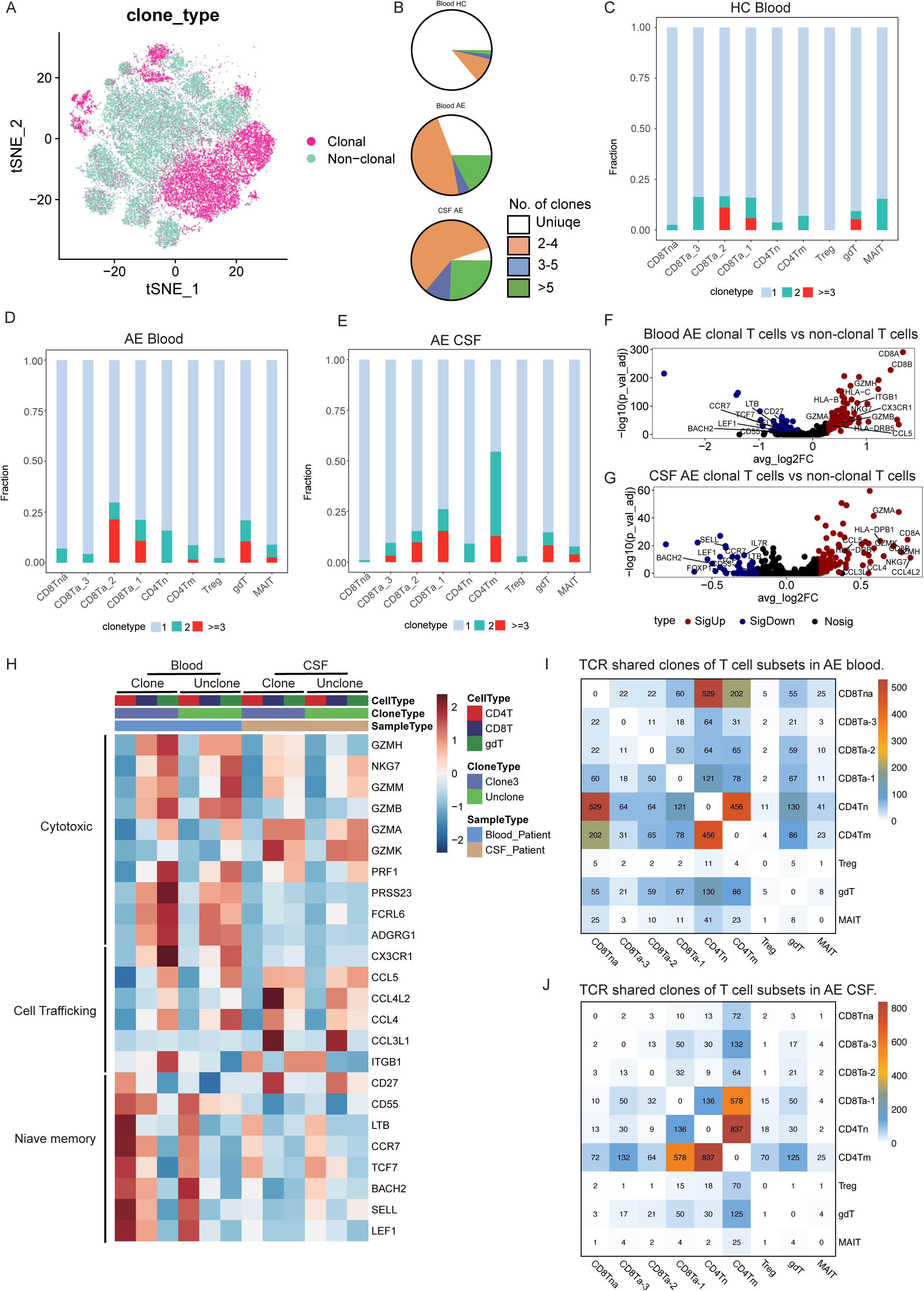
Significant TCR clone expansion of CD8^+^T cells in both CSF and PB of NMDAR-E patients. (A) Distribution of clonal cells in T cell clusters found in the NMDAR-E patients (CSF, n=3; PB, n=4) and healthy controls (HC) (n=2). (B) TCRαβ clonality of T cells in CSF and PB from NMDAR-E patients, as well as blood samples from HC, represented with clones color-coded according to their proportion of total TCRαβ sequences. (C-E) Bar graphs illustrating the cellular proportions of three different clonal expansion groups in each T cell cluster, derived from PB of HC (C), PB of NMDAR-E patients (D), and CSF of NMDAR-E patients (E). (F-G) Volcano plots comparing the differential gene expression (DEGs) between highly TCR clonal (clonal TCRαβ ≥ 3) and unclonal T cells in both blood (F) and CSF (G) from NMDAR-E patients. Detailed source data for (F, G) are provided in Table S9. (H) Heatmaps showing selected DEGs in highly clonal (clonal TCRαβ ≥ 3) in PB and CSF of NMDAR-E patients, compared to unclonal T cells. Heatmap colors indicate z-scored values for mean gene expression in each group. (I-J) Sharing of expanded TCR clonotypes across T cell clusters in the PB (I) and CSF (J) of NMDAR-E patients. The bottom heatmap shows the number of shared expanded TCR clonotypes for each cluster pair in NMDAR-E patients.

### 3.4 In vitro co-cultured of CD8^+^T and B cells

To explore the interaction between CD8^+^T cells and B cells in NMDAR-E, we isolated PBMCs from both NMDAR-E patients and control individuals, and sorted CD8^+^T cells and CD19^+^B cells for co-culture. On the first day of co-culture, flow cytometry analysis indicated no significant difference in the apoptosis rate of CD8^+^T and B cells between the control group and the NMDAR-E group (Figure 4A, 4B and 4D). However, on the 7th day of co-culture, apoptosis detection revealed a significantly higher apoptosis rate of B cells in the NMDAR-E group compared to the control group, while the apoptosis rate of CD8^+^T cells showed no significant difference between them (Figure 4C and 4E). Specifically, the apoptosis proportions of antibody-secreting cells (ASCs), memory B cells (Bm), CD27^-^ B cells, and CD27^-^IgD^-^ double negative B (DN) cells in the NMDAR-E group were significantly higher than those in the control group (Figure 4F-4I). These results suggest a potential interaction between CD8^+^T cells and B cells in NMDAR-E patients, leading to the apoptosis of B cell subpopulations.

**Figure 4.**
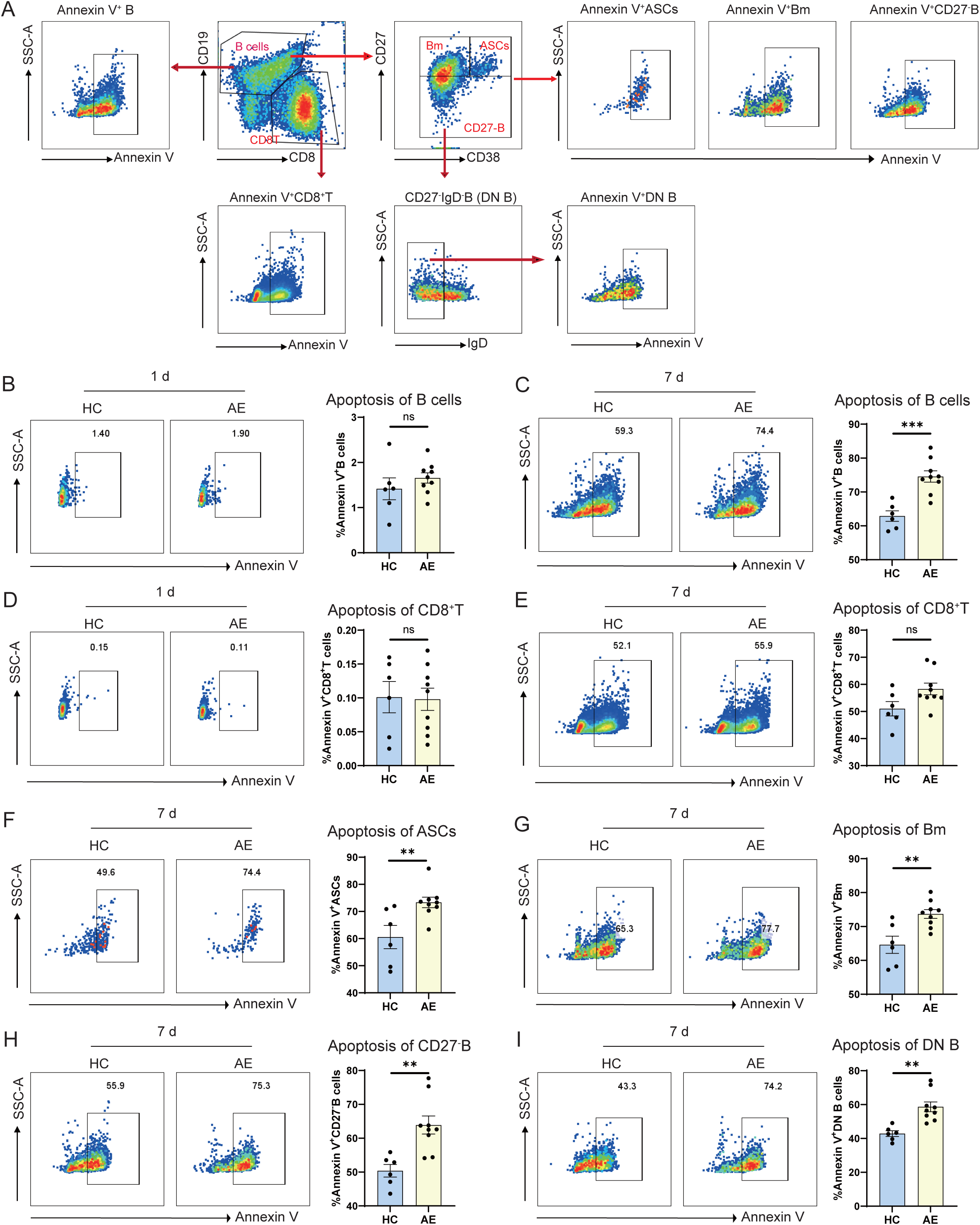
In vitro co-culture of B cells and CD8^+^T cells from NMDAR-E patients and controls. (A) Flow charts of flow cytometry. (B-E) Representative contour plots and summary scatter plots showing the percentage of annexin V^+^ B cells (B-C) and annexin V^+^ CD8^+^T cells (D-E) at 1 day and 7 day of co-culture from PB of 6 healthy controls (HC) and 9 NMDAR-E patients. (F-I) Representative contour plots and summary scatter plots showing the percentage of annexin V^+^ASCs, annexin V^+^ memory B cells (Bm), annexin V^+^ CD27-, annexin V^+^ DN cells at 7 day of co-culture from the PB of HCs and NMDAR-E patients. * < 0.05; **P < 0.01; ***P < 0.001; evaluated using two-tailed unpaired Student’s t test.

### 3.5 Alteration of KIR^+^CD8^+^T cells in NMDAR-E

Previous research has indicated the presence of a regulatory subset of KIR^+^CD8^+^T cells within the CD8^+^T cell population, characterized by high expression of Killer cell immunoglobulin-like receptor (KIR) genes. To explore this CD8^+^T cell subsets, we analyzed the expression of KIRs in CD8^+^T cells. The results revealed higher expression of *KIR3DL1* and *KIR2DL3* in CD8^+^T cells in the CSF and PB of NMDAR-E patients compared to the control group, and these KIR^+^CD8^+^T cells were predominantly composed of activated CD8^+^T cells (Figure 5A-5C). Further flow cytometry analysis of KIR^+^CD8^+^T cells in the PB of NMDAR-E patients and the control group revealed a higher level of CD158b^-^CD158e^+^CD8^+^T (KIR^+^CD8^+^T) cells in the severe group compared to the control group (Figure 5D and 5E). Additionally, we observed a significant increase in the level of CD45RA^+^CCR7^-^CD158b^-^ CD158e^+^CD8^+^T (KIR^+^CD8^+^Temra) cells in relapse-phase NMDAR-E patients (Figure 5F).

**Figure 5.**
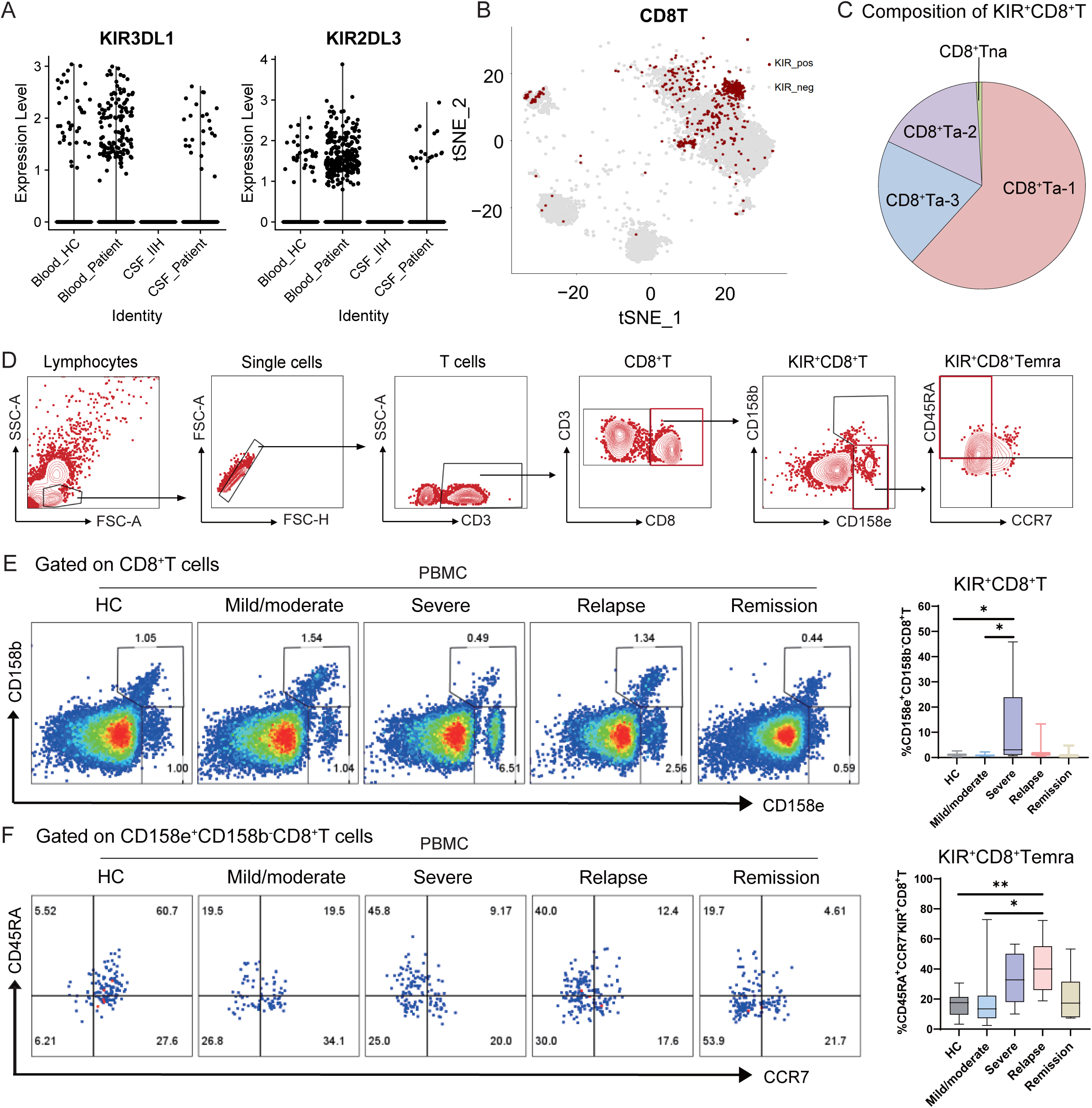
Changes of KIR^+^CD8 T cells in NMDAR-E patients. (A) Expression of KIR transcripts (KIR3DL1 and KIR2DL3) in CD8^+^T cells from the CSF and PB of control individuals (CSF, n= 6; Blood, n=2) and NMDAR-E patients (CSF, n=3; Blood, n=4). (B) t-SNE plots showing the distribution of KIR^+^CD8^+^T cells (expressing KIR3DL1 and/or KIR2DL3 genes) and KIR^-^CD8^+^T cells in the CSF and PB of controls and NMDAR-E patients. (C) The composition of KIR^+^CD8^+^T cell population shown in Fig.5B. (D) Flow charts of KIR^+^CD8^+^T cell subclusters. (E-F) Representative contour plots and summary scatter plots show the percentage of CD158e^+^cells in CD8^+^T cells (E) and CD45RA^+^cells in CD158e^+^CD8^+^T cells (KIR^+^CD8^+^Temra) (F) from the PB of HC (n=11) and NMDAR-E patients (mild/moderate group, n=10; severe group, n=9; relapsed group, n=13; remission group, n=8).

### 3.6 TCR clonal characteristics of KIR^+^CD8^+^T and their interaction with B cells

scTCR-seq analysis revealed clonal expansion of KIR^+^CD8^+^T cells in both the CSF and PB of NMDAR-E patients (Figure 6A and 6B). Compared to KIR^+^CD8^+^T cells without TCR clonality, those with TCR clonality in the CSF and PB of NMDAR-E patients upregulated DEGs associated with NK cells (such as *KLRF1*, *KLRD1*, *KLRB1*) and were enriched in the natural killer cell-mediated cytotoxicity pathways (Figure 6C-6G). Additionally, KIR^+^CD8^+^T cells with TCR clonality in the PB of NMDAR-E patients upregulated genes related to cytotoxicity while downregulating genes associated with naive/memory markers (Figure 6C and 6E).

**Figure 6.**
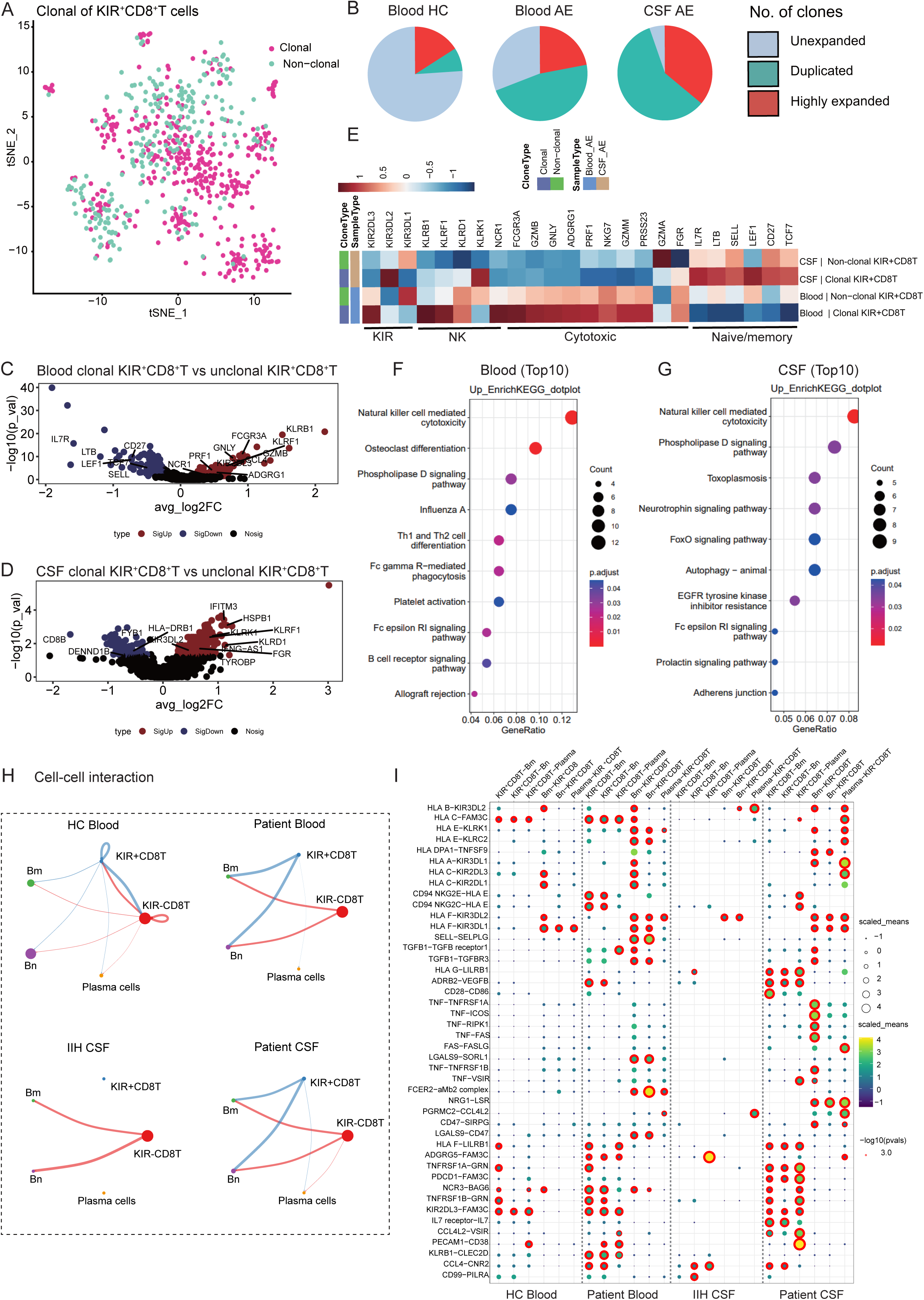
TCR clonal characteristics of KIR^+^CD8^+^T and their interaction with B cells. (A) t-SNE plots of KIR^+^ CD8^+^T cells from NMDAR-E patients and controls with clonal and unclonal cells annotated with different colors (clonal, red; unclonal, green). (B) TCRαβ clonality of KIR^+^CD8^+^T in the CSF and PB of NMDAR-E patients and PB of healthy control (HC). Clones are colored by proportion of the total TCRαβ sequences. (C-D) The differential gene expression (DEG) of expanded TCR clones revealed increased expression of cytotoxic effector genes in the clonal KIR^+^CD8^+^T cells of NMDAR-E patients as compared to unclonal KIR^+^CD8^+^T in both PB (C) and CSF (D) samples. Detailed source data for (C, D) are provided in Table S10. (E-F) Analysis of the top 10 KEGG pathways enriched for DEGs in KIR^+^CD8^+^T cells with and without TCR clones in the PB (E) and CSF (F) of NMDAR-E patients. (H) Cell-cell interaction analysis predicts interactions between KIR^+^CD8^+^T cells and KIR^-^ CD8^+^T cells with plasma cells, naïve B (Bn) cells, and memory B (Bm) cells in the CSF and PB of controls and NMDAR-E patients. (I) Dot plot illustrating cell-cell interactions between KIR^+^CD8^+^T cells and plasma cells, Bn cells, and Bm cells based on selected cytokines and molecules calculated by CellPhoneDB.

Cell interaction analysis indicated stronger interactions between KIR^+^CD8^+^T cells from the CSF and PB of NMDAR-E patients with naive B cells (Bn) and memory B cells (Bm) compared to the control group (Figure 6H). Further receptor-ligand pairing analysis demonstrated interactions between KIR genes (including *KIR3DL1*, *KIR2DL3*, and *KIR2DL1*) and NK-related molecules (*KLRC2* and *KLRK1*) expressed on KIR^+^CD8^+^T cells from the CSF and PB of NMDAR-E patients, and MHC-I molecules (including *HLA-A*, *HLA-C*, and *HLA-E*) expressed on B cells (Figure 6I).

## 4. Discussion

This study used scRNA-seq, scTCR-seq, and flow cytometry to analyze changes in T cell subpopulations, transcriptional features, and immune characteristics in NMDAR-E patients for the first time. Additionally, we observed that CD8^+^T cells from NMDAR-E patients promoted apoptosis of their own B cells, providing direct evidence for the connection between CD8^+^T cells and B cells, and offering new insights into the role of T cell subsets in the pathogenesis of NMDAR-E.

Despite previous research on NMDAR-E primarily focusing on the roles of B cells and autoantibodies, there is increasing attention on the role of T cells in autoimmune encephalitis ^41^. The cytokine/chemokine profile of human NMDAR-E cases in CSF supports evidence for T cell involvement ^14, 15, 42, 43^. Currently, only a few studies have reported histopathological changes in NMDAR-E patients, with some cases showing immune cell infiltration and neuroinflammation ^9, 17, 44, 45^. For example, one report noted the absence of plasma cells and B cells in brain tissue but persistent infiltration of CD4^+^ and CD8^+^ T cells following ineffective treatment with plasma exchange and rituximab ^44^. Another report described NMDAR antibody-positive paraneoplastic encephalitis associated with SCLC, where cytotoxic T cells positive for CD3, CD8, TIA-1, and granzyme B were widely distributed in brain parenchyma ^45^. However, there are also reports where T cell infiltration in brain tissue was not prominent ^18^, possibly influenced by timing and location of sample collection, reflecting individual variability. Although NMDAR-E mouse models show significantly higher average infiltration density of CD4^+^ and CD8^+^ T cells in the hippocampus six weeks post-immunization compared to controls ^20^, studies using antigen-reactive T cell enrichment techniques have found lower frequencies of CD154-expressing NR1-reactive CD4^+^ helper T cells in NMDAR-E patients compared to healthy controls ^46^. Therefore, research on T cell subsets in NMDAR-E remains limited, and their exact roles and alterations in disease mechanisms require further investigation. This study employed scRNA-seq and flow cytometry analysis, revealing no significant overall changes in the proportion of CD4^+^ and CD8^+^ T cells in CSF and PB of NMDAR-E patients compared to controls, but a significantly higher proportion of activated CD8^+^T cell subsets, particularly CD8^+^Temra cells, in recurrent NMDAR-E patients’ PB compared to controls and acute phase group. These findings suggest that beyond overall CD8^+^T cell infiltration, specific CD8^+^T cell subsets and their transcriptomic characteristics warrant further exploration to elucidate their roles in NMDAR-E.

After understanding the changes in T cell subpopulations in the CSF and PB of NMDAR-E patients, this study analyzed the transcriptional features of different T cell subpopulations using scRNA-seq. We observed that activated CD8^+^T cell subpopulations (such as CD8^+^Ta_1 and CD8^+^Ta_2) in NMDAR-E patient CSF, as well as CD4^+^Tm subpopulations, expressed tissue-resident related genes like *CD69* and *CXCR6*. *CD69*, a major phenotypic marker of tissue-resident memory T cells (Trm), is a transmembrane glycoprotein that can influence T cell migration by downregulating S1PR1 expression ^47^. Trm cells have the ability to secrete pro-inflammatory cytokines and attract other leukocytes ^48^. Additionally, *CXCR6* is also a characteristic marker of Trm cells ^49, 50^. In the CNS, the CXCL16/CXCR6 signaling pathway regulates the number of CD8^+^Trm cells and plays a role in their interaction with microglial cells ^49^. Furthermore, DEGs showed higher expression of *CXCR3* (the receptor for chemokines CXCL9 and CXCL10) in CD4^+^ and CD8^+^T cell subpopulations in NMDAR-E patient CSF. *CXCR3* plays a crucial role in the development of Trm cells ^51^. Several studies have reported an increase in CXCL10 cytokine levels in the CSF of NMDAR-E patients ^15, 16, 52^. Therefore, the tissue-resident characteristics expressed by T cell subpopulations in NMDAR-E may influence their migratory capabilities or recruit other leukocytes (such as B cell subpopulations) to infiltrate the CNS through the secretion of inflammatory cytokines.

In addition, we observed that T cell subsets (including CD4^+^Tm, CD8^+^Ta, and gdT cells) in the CSF and PB of NMDAR-E patients exhibit elevated expression of characteristic genes associated with TH1 cells, such as *TBX21* and *CXCR3*. TH1 cells can secrete cytokines like IFN-γ, TNF-α, IL-2, GM-CSF, promoting T cell proliferation, differentiation, and maturation ^53^. Previous studies have also reported significant elevation of TH1 axis cytokines in the CSF of NMDAR-E patients, including IFN-γ ^15^, CXCL10 ^15, 16, 52^, and TNF-α ^15, 52, 54^. Furthermore, T cell subsets in the CSF of NMDAR-E patients show upregulation of IFN-related genes, including *IFNGR1*, *IRF7*, and *STAT1*. *IRF7*, as a member of the interferon regulatory factor family, activates the JAK-STAT pathway, inducing the expression of type I interferon genes, thereby enhancing the production of antiviral proteins, demonstrating robust antiviral functionality ^55^. Type II interferon (IFN-γ), upon binding with its receptor complex (IFNGR1 and IFNGR2), primarily stimulates the maturation and differentiation of immune cells via STAT1 activation, thereby enhancing adaptive immune responses, demonstrating superior immune regulatory effects ^56^. In summary, activation of the TH1 and IFN-related pathways in T cells of NMDAR-E patients may promote the maturation and differentiation of immune cells, playing a potential role in immune regulation.

Upon recognition of homologous antigens, CD8^+^T cells undergo rapid clonal expansion and differentiate into effector and memory cells ^57^. Our study identified some subsets of effector CD8^+^T and CD4^+^Tm cells in the CSF and PB of NMDAR-E patients exhibiting TCR clonality. DEGs analysis revealed that these CD8^+^T cells with TCR clonality possess potential cytotoxic and chemotactic properties. Studies in CNS diseases such as multiple sclerosis ^58^, Parkinson’s disease ^59^, and Alzheimer’s disease ^39^ have also reported TCR expansions. For instance, in Alzheimer’s disease, autoreactive cytotoxic T cells can recognize self-antigens, leading to target cell death and exacerbating neuroinflammation ^39^. Further investigation into the role of autoreactive CD8^+^T cells in CNS diseases is warranted. For example, identifying TCR sequences specific to NMDA autoantigens could enhance our understanding of the pathogenesis of NMDAR-E and lay the groundwork for developing new diagnostic and therapeutic approaches.

Interesting findings from this study revealed that CD8^+^T cells from NMDAR-E patients can promote apoptosis of their own B cells, suggesting an interaction between CD8^+^T cells and B cells in NMDAR-E encephalitis. Although traditionally most CD8^+^T cells are thought to be involved in controlling infections caused by pathogens or targeting cancer cells, emerging evidence suggests that there is a subset of CD8^+^T cells with the function of regulating the self-immune response^60^. Human KIR^+^CD8^+^T cells function similarly to the immune-suppressive subset Ly49^+^CD8^+^T cells in mice, increasing in certain autoimmune or infectious diseases (e.g., celiac disease, multiple sclerosis, COVID-19), and maintaining immune tolerance by suppressing excessive activation of immune cells ^40, 61-63^. In NMDAR-E, the cytotoxic effect of CD8^+^T cells on self-B cells may be attributed to these regulatory CD8^+^T cell subsets, which can mediate B cell apoptosis. This study found that the transcriptional characteristics of KIR^+^CD8^+^T cells in NMDAR-E are consistent with the findings of Li et al.^40^. Cell interaction analysis indicated that B cells in the CSF and PB of NMDAR-E patients express various MHC-I molecules, which interact with KIR^+^CD8^+^T cells through receptor-ligand interactions. Li et al. demonstrated that human KIR^+^ CD8^+^T cells can kill pathogenic CD4^+^T cells via MHC-I molecules ^40^. Similarly, mouse Ly49^+^ CD8^+^T cells depend on perforin and classical/non-classical MHC-I molecules ^64^. Therefore, there may exist a subset of CD8^+^T cells in NMDAR-E that regulate immune responses by modulating the cytotoxicity against self-B cells, although the specific mechanisms require further exploration.

Our study has several limitations. Firstly, we used publicly available databases as controls for CSF, which may introduce potential batch effects. To mitigate this, we employed scVI methods for data analysis to enhance the reliability and validity of our findings. Secondly, cell communication analysis revealed interactions between KIR^+^CD8^+^T cells and B cell subsets. However, due to the very low numbers of KIR^+^CD8^+^T cells in clinical samples, it was challenging to isolate a sufficient quantity from CSF or PBMCs for further study. Lastly, our study samples were derived from NMDAR-E patients’ CSF and PB. While this method reflects the actual disease state directly, it also limits the depth of our investigation. Future research could involve establishing an NMDAR-E mouse model to delve deeper into the roles and mechanisms of T cell subsets in NMDAR-E.

## Conclusion

This study found that the proportion of activated CD8^+^T cell subsets in the CSF and PB of NMDAR-E patients is elevated, accompanied by TCR clonality. Differential gene expression analysis revealed upregulation of genes related to cytotoxicity, tissue residency, Th1, IFN, or TCR signaling in some activated CD8^+^T cell and CD4^+^T memory cell subsets. In vitro co-culture experiments demonstrated that CD8^+^T cells from PB of NMDAR-E patients can promote apoptosis of their own B cells. Further cell interaction analysis revealed interactions between KIR^+^CD8^+^T cells and B cell subsets in NMDAR-E patients. However, whether KIR^+^CD8^+^T cells mediate apoptosis of their own B cells in NMDAR-E requires further exploration. Overall, this study suggests that CD8^+^T cell subsets may participate in the pathogenesis of NMDAR-E by influencing B cell subsets, offering potential value for exploring new therapeutic approaches in the future.

## Acknowledgment

This work was supported by grants from Chongqing Natural Science Foundation (Grant No. 2024NSCQ-MSX0894), National Science Foundation of China (Grant No. 81571272 and 82071459), and the 1.3.5 project for disciplines of excellence and Brain Science project of West China Hospital, Sichuan University (Grant No. ZYJC21001). The authors express their gratitude to Genenergy Shanghai for their support, as well as to all the patients who generously participated in the study. Thanks to Dr. Yuan Liang for his help in our data analysis.

## Author contributions

JL and DZ designed this study and revised the final version of the draft. SL and XH carried out this study; those authors collected samples, analyzed data, wrote the draft, and contributed equally to this study; YY, and JW assisted in collecting the samples; ZH edited the manuscript.

## Data availability statement

The raw sequence data reported in this article are currently being uploaded to the Genome Sequence Archive (GSA: PRJCA014916) at the National Genomics Data Center, Beijing Institute of Genomics, Chinese Academy of Sciences. Once the upload is complete, these data will be publicly accessible at https://ngdc.cncb.ac.cn/gsa. Currently, interested parties can request access to the relevant data by contacting the corresponding author. The data sets supporting the conclusions of this article are included within the article and its accompanying files.

## Supplemental information

**Document S1. Figures S1-S2 and Table S1-S6**

**Table S7.** Differential expression genes of CD4^+^T and CD8^+^T cells in the CSF of NMDAR-E patients versus IIH.

**Table S8.** Differential expression genes of CD4^+^T and CD8^+^T in the PB of NMDAR-E patients versus healthy controls.

**Table S9.** Differential expression genes of clonal T cells versus unclonal T cells from NMDAR-E patients in both blood and CSF samples.

**Table S10.** Differential expression genes of clonal KIR^+^CD8^+^T cells versus unclonal KIR^+^CD8^+^T cells from NMDAR-E patients in both blood and CSF samples.

**Table S7-S10 are provided in separate Excel spreadsheets.**

## Abbreviations

NMDAR: Anti-N-methyl-D-aspartate receptor
AE: autoimmune anti-NMDAR encephalitis
CBA: cell-based assay
CNS: central nervous system
CSF: cerebrospinal fluid
PBMCs: Peripheral blood mononuclear cells
PB: peripheral blood
scTCR-Seq: single-cell TCR sequencing
scRNA-seq: Single cell RNA sequencing
Temra: CD45RA^+^ effector memory T cells
Tem: effector memory T
Tcm: central memory T
Tn: naïve T
HC: Healthy control
IIH: Idiopathic intracranial hypertension
Bm: memory B cells
Bn: naïve B cells
ASCs: Antibody-secreting cells
DN: CD27-IgD-double negative B cells
t-SNE: t-distributed neighbor embedding
MHC: Major histocompatibility complex
DEGs: Differentially expressed genes
KIRs: killer-cell immunoglobulin-like receptors
Trm: tissue-resident memory T cells.

