## Supplementary material for "Single-cell analyses of CSF and PBMCs from anti-NMDAR encephalitis patients reveals distinct characteristics of T cell subpopulations": Document S1

**Table S1. Demographic information of patients with NMDAR-E and controls included for single cell RNA sequencing (scRNA-seq) and single cell TCR sequencing (scTCR-seq).**

| Patient  /Control | scRNA-seq and scTCR-seq | Sample | Age Range | Sex | Disease Progression at Sample Collection | NMDAR antibodies titer (CSF/Serum) | Primary clinical manifestations |
| --- | --- | --- | --- | --- | --- | --- | --- |
| AE1 | Both | PB | 21-25 | F | 17d | 1:1000/1:1000 | Epilepsy, mental disorders, impaired consciousness |
| AE2 | Both | PB | 26-30 | M | 14d | 1:100/1:1000 | Headache, epilepsy, mental disorders, impaired consciousness |
| AE3 | Both | PB | 21-25 | F | 14d | 1:1000/1:1000 | Epilepsy, mental disorders, impaired consciousness |
| AE4 | Both | PB | 31-35 | F | 26d | 1:100/1:100 | Mental disorders, cognitive impairments |
| HC1 | Both | PB | 26-30 | M | NA | NA | NA |
| HC2 | Both | PB | 21-25 | F | NA | NA | NA |
| AE5 | Both | CSF | 36-40 | F | 2m (Relapse 3d) | 1:100/1:100 | Mental disorders, cognitive impairments |
| AE6 | Both | CSF | 61-65 | M | 20d | 1:100/1:100 | Headache, fever, and mental disorders, epilepsy |
| AE7 | Both | CSF | 16-20 | F | 17d | 1:100/1:100 | Impaired consciousness and mental disorders |
| IIH1 | scRNA-seq | CSF | 41-45 | F | NA | NA | NA |
| IIH2 | scRNA-seq | CSF | 41-45 | M | NA | NA | NA |
| IIH3 | scRNA-seq | CSF | 31-35 | F | NA | NA | NA |
| IIH4 | scRNA-seq | CSF | 21-25 | F | NA | NA | NA |
| IIH5 | scRNA-seq | CSF | 31-35 | M | NA | NA | NA |
| IIH6 | scRNA-seq | CSF | 21-25 | F | NA | NA | NA |

The data of IIH came from public dataset GSE138266. For scRNA-seq, samples AE1 and AE2, AE3 and AE4, as well as HC1 and HC2 were combined together respectively. AE, anti-NMDAR autoimmune encephalitis; IIH, idiopathic intracranial hypertension; HC, healthy control; CSF, cerebrospinal fluid; PB, peripheral blood; F, female; M, male; d, day; m, month; NA, Not Applicable.

**Table S2. Demographic characteristics of patients with NMDAR-E and control individuals undergoing flow cytometry analysis of T cell subpopulations in CSF.**

|  | NMDAR-E | Controls |
| --- | --- | --- |
| Number | 15 (55.6%) | 12 (44.4%) |
| Sex |  |  |
| Male | 8 (53.3%) | 7 (58.3%) |
| Female | 7 (46.7%) | 5 (41.7%) |
| Age (years, mean±SEM) | 29.26±9.25 | 30.9±10.9 |
| Duration of the disease at the time of sampling |  |  |
| ≤1 month | 5 (33.3%) | NA |
| 1-3 month | 7 (46.7%) | NA |
| >3month | 3 (20.0%) | NA |
| Antibody titer in CSF |  |  |
| ≤1:10 | 4 (26.7%) | NA |
| 1:10-1:100 | 9 (60.0%) | NA |
| >1:100 | 2 (13.3%) | NA |
| Antibody titer in serum |  |  |
| ≤1:10 | 3 (20.0%) | NA |
| 1:10-1:100 | 8 (53.3%) | NA |
| >1:100 | 4 (26.7%) | NA |
| mRS score |  |  |
| 1-2 | 5 (33.3%) | NA |
| 3 | 3 (20.0%) | NA |
| 4-5 | 7 (46.7%) | NA |

mRS, Modified Rankin Scale; CSF, cerebrospinal fluid; NA, Not Applicable.

**Table S3. Demographic information of NMDAR-E patients and control individuals who underwent flow cytometry analysis of T cell subpopulations in PB.**

|  | Acute phase of NMDAR-E | Relapse phase of NMDAR-E | Controls |
| --- | --- | --- | --- |
| Number | 34 (50%) | 13(19.1) | 21 (30.9%) |
| Sex |  |  |  |
| Male | 18 (52.9%) | 6 (46.2%) | 12 (57.1%) |
| Female | 16 (47.1%) | 7 (53.8%) | 9 (42.9 %) |
| Age (years, mean±SEM) | 31.76±11.56 | 37.8±18.4 | 26.57±3.09 |
| Duration of the disease at the time of sampling |  |  |  |
| ≤1 month | 21 (61.8%) | 0 (0%) | NA |
| 1-3 month | 13 (38.2%) | 0 (0%) | NA |
| 3-12 month | 0 (0%) | 4 (30.8%) | NA |
| 1-2 year | 0 (0%) | 4 (30.8%) | NA |
| 2-5 year | 0 (0%) | 5 (38.5%) | NA |
| Antibody titer in CSF |  |  |  |
| ≤1:10 | 12 (35.3%) | 6 (46.2%) | NA |
| 1:10-1:100 | 18 (52.9%) | 7 (53.8%) | NA |
| >1:100 | 4 (11.8%) | 0 (0%) | NA |
| Antibody titer in serum |  |  |  |
| ≤1:10 | 9 (26.5%) | 3 (23.1%) | NA |
| 1:10-1:100 | 17 (50.0%) | 7 (53.8%) | NA |
| >1:100 | 8 (23.5%) | 3 (23.1%) | NA |
| Disease severity |  |  |  |
| Mild/moderate (mRS≤3) | 13 (38.2%) | 11 (84.6%) | NA |
| Severe (mRS >3) | 21 (61.8%) | 2 (15.4%) | NA |
| Seizure | 28 (82.3%) | 7 (53.8%) | NA |
| Status epilepticus | 9 (26.5%) | 1 (7.8%) | NA |
| Mental disorder | 31 (91.2%) | 10 (76.9%) | NA |
| Impaired consciousness | 26 (76.5%) | 10 (76.9%) | NA |

mRS, Modified Rankin Scale; CSF, cerebrospinal fluid; NMDAR-E, anti-NMDAR encephalitis; NA, Not Applicable.

**Table S4. Demographics information of NMDAR-E patients and control individuals who underwent flow cytometry analysis of KIR^+^CD8^+^T cell subpopulations in periphery blood.**

|  | Acute-phase (severe) | Acute-phase (mild/moderate) | Relapse-phase | Remission-phase | Controls |
| --- | --- | --- | --- | --- | --- |
| Number | 9 (17.6%) | 10 (19.6%) | 13 (25.5%) | 8 (15.7%) | 11 (21.6%) |
| Sex |  |  |  |  |  |
| Male | 6 (66.7%) | 5 (50.0%) | 6 (46.2%) | 5 (62.5%) | 7 (63.6%) |
| Female | 3 (33.3%) | 5 (50.0%) | 7 (53.8%) | 3 (37.5%) | 5 (45.5%) |
| Age (years, mean±SEM) | 39.8±14.0 | 29.7±12.4 | 37.8±18.4 | 26.6±12.0 | 27.18±3.28 |

**Table S5. The specific information for flow cytometry antibodies in the study.**

| ANTIBODIES | SOURCE | IDENTIFIER |
| --- | --- | --- |
| Flow Cytometry: rat anti-human CD16/CD32 Fc Block | BioLegend | [Cat# 422301](http://admin.bioec.cn/index.php?r=product/view&id=94958" \o "http://admin.bioec.cn/index.php?r=product/view&id=94958) |
| Flow Cytometry: anti-human CD3-BV605 | BioLegend | Cat# 317321 |
| Flow Cytometry: anti-human CD8-APC-Cy7 | BioLegend | Cat# 301016 |
| Flow Cytometry: anti-human CD4-PerCP-Cy5-5 | BioLegend | [Cat# 317427](http://admin.bioec.cn/index.php?r=product/view&id=173332" \o "http://admin.bioec.cn/index.php?r=product/view&id=173332) |
| Flow Cytometry: anti-human CCR7-FITC | BioLegend | Cat# 353216 |
| Flow Cytometry: anti-human CD45RA -BV786 | BioLegend | Cat# 304139 |
| Flow Cytometry: anti-human CD158b-PerCP-Cy5-5 | BioLegend | Cat#312614 |
| Flow Cytometry: anti-human CD158e-PE | BioLegend | Cat# 312708 |
| Flow Cytometry: anti-human CD19-BV785 | BioLegend | Cat# 363028 |
| Flow Cytometry: anti-human CD38-APC | BioLegend | Cat#356605 |
| Flow Cytometry: anti-human CD27-PE-Cy7 | BioLegend | Cat# 356411 |
| Flow Cytometry: anti-human IgD-APC-Cy7 | BioLegend | Cat# 348217 |
| Flow Cytometry: AnnexinV-PE | BD Pharmingen | Cat#559763 |
| Flow Cytometry: 7AAD-PerCP-Cy5-5 | BioLegend | Cat# 420403 |

**Table S6. Characteristics of anti-NMDAR encephalitis patients and control individuals for single cell RNA sequencing.**

| Group | Sample | Number of cells | Median UMI counts per cell | Median genes per cell | Number of cells passing QC |
| --- | --- | --- | --- | --- | --- |
| Patient blood | PBMC | 20105 | 3432 | 1477 | 19159 |
| HC blood | PBMC | 7761 | 2241 | 960 | 7508 |
| Patient CSF | CSF | 19432 | 2997 | 1376 | 18421 |
| IIH CSF | CSF | 17421 | 2639 | 782 | 14536 |

The data of IIH came from public dataset GSE138266. AE, Autoimmune encephalitis; IIH, idiopathic intracranial hypertension; HC, healthy control.

2. **Figures**

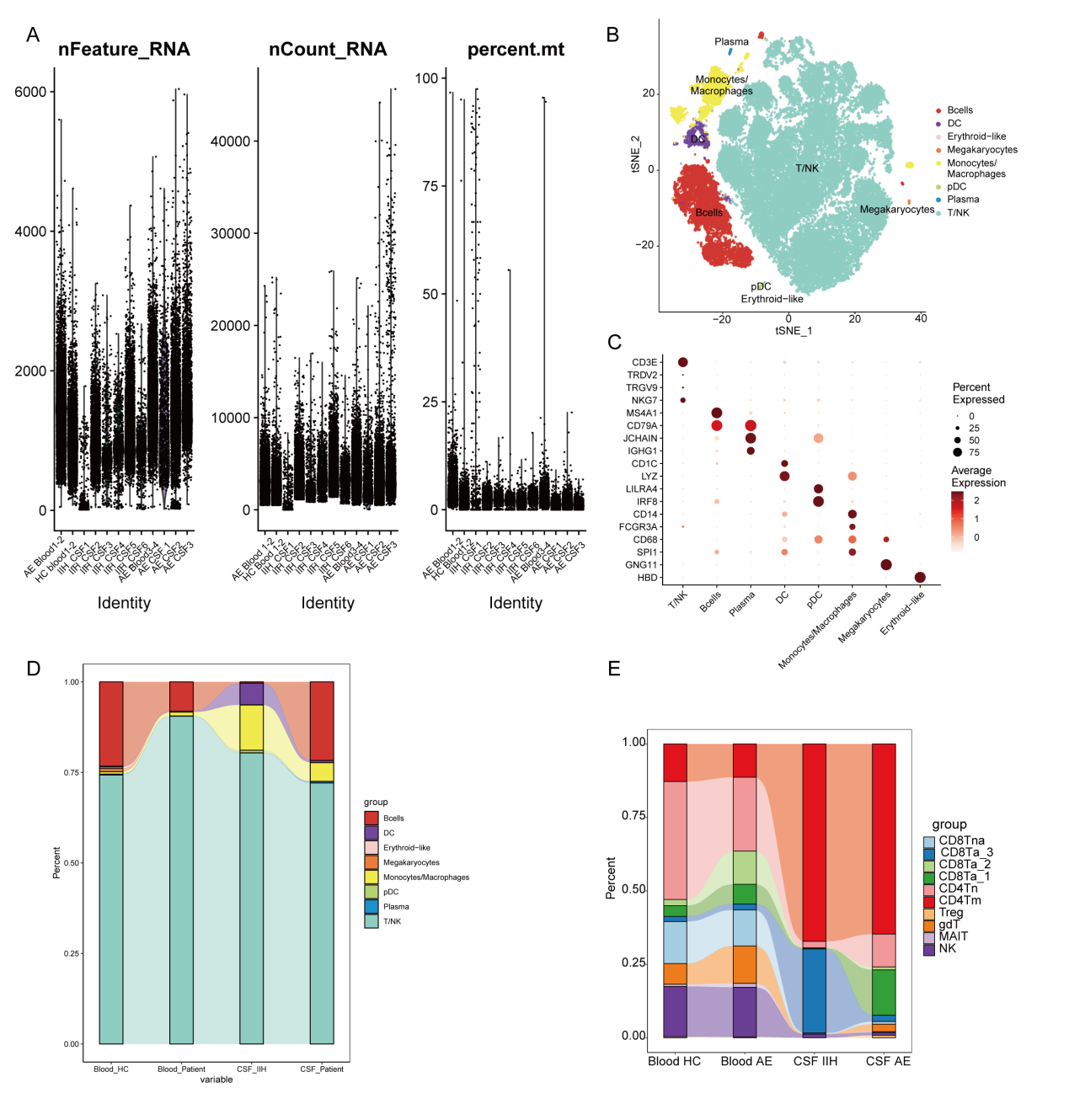

**Figure S1. Single cell transcriptomic analysis of immune cells across CSF and blood in patients with anti-NMDAR encephalitis (NMDAR-E) and control individuals.** (A) This study included quality control measures for samples undergoing single cell RNA sequencing analysis. (B) t-SNE plot showing the distribution of immune cell subsets in blood and CSF in control individuals and NMDAR-E patients. (C) Marker gene expression used to classify subclusters of all cells among CSF and blood. (D) The bar chart illustrates the proportion of immune cell subtypes in the cerebrospinal fluid and peripheral blood of NMDAR-E patients and control groups. (E) The bar chart illustrates the proportion of T/NK cell subtypes in the CSF and PB of NMDAR-E patients and control groups.

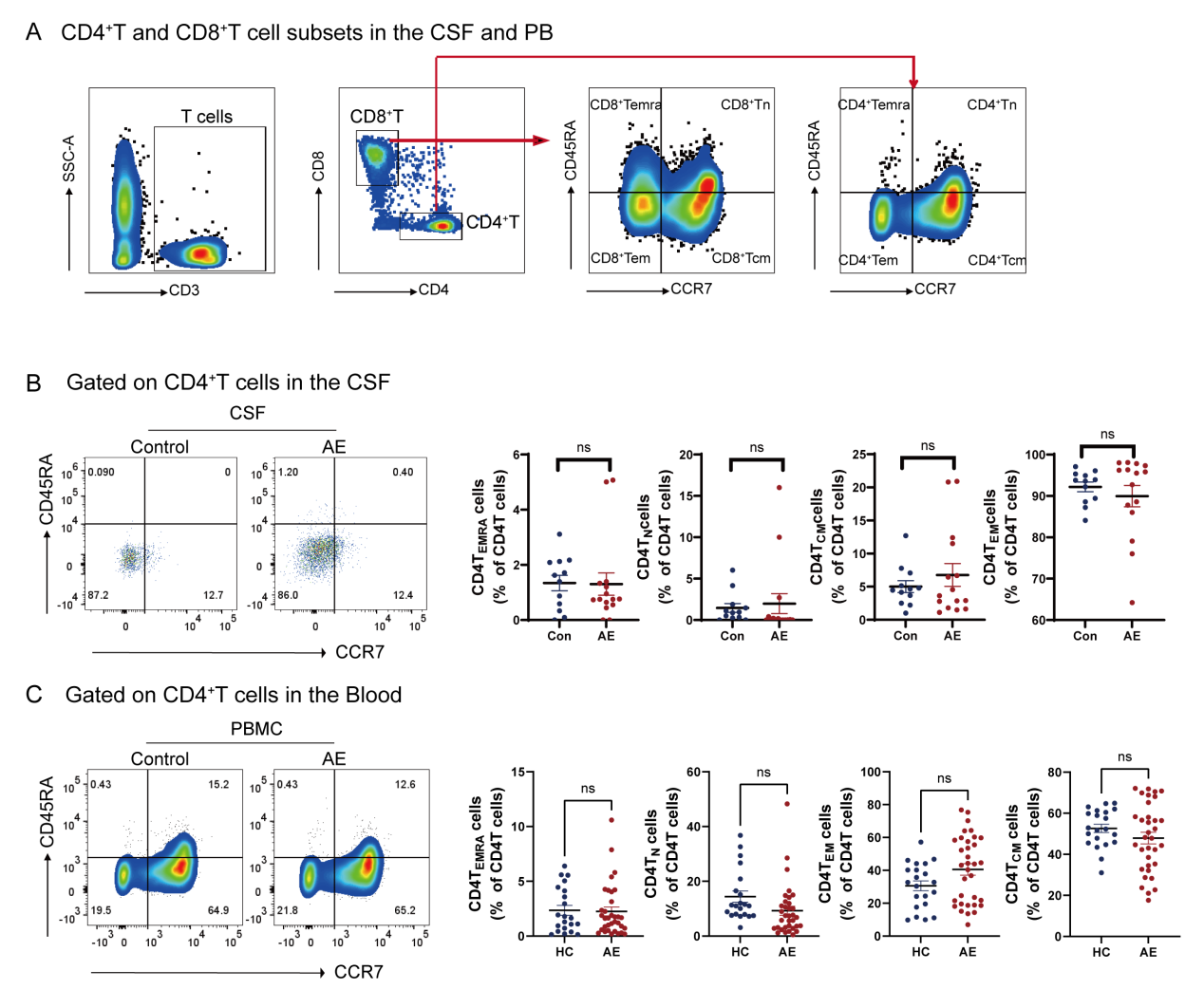

**Figure S2. Analysis of T cell subset changes by flow cytometry.** (A) Flow charts of CD8^+^T and CD4^+^T cell subsets in the CSF and PB of NMDAR-E patients and controls; (B-C) Flow cytometry was used to detect the changes of CD4^+^T subsets in CSF (B) and PB (C) of NDMAR-E patients and controls.

**Table S7.** Differential expression genes of CD4^+^T and CD8^+^T cells in the CSF of NMDAR-E patients versus IIH.

**Table S8.** Differential expression genes of CD4^+^T and CD8^+^T in the PB of NMDAR-E patients versus healthy controls.

**Table S9.** Differential expression genes of clonal T cells versus unclonal T cells from NMDAR-E patients in both blood and CSF samples.

**Table S10.** Differential expression genes of clonal KIR^+^CD8^+^T cells versus unclonal KIR^+^CD8^+^T cells from NMDAR-E patients in both blood and CSF samples.

**Table S7-S10 are provided in separate Excel spreadsheets.**
